# Safety and Immunogenicity of a Booster SARS-Cov-2 Vaccination in Patients with Chronic Liver Disease

**DOI:** 10.1101/2022.10.30.22281713

**Authors:** Li Qianqian, Chen Jing, Sun Kai, Song Ruixin, Wang Jiayin, Lv Hongmin, Yang Yankai, Liang Jing, Ye Qing, Gao YanYing, Li Jun, Li Ying, Yan Junqing, Yang Chao, Liu Ying, Wang Tao, Liu Changen, Wang Fei, Xiang Huiling

## Abstract

This article is aim to investigate the safety and immunogenicity of the severe acute respiratory syndrome coronavirus 2 (SARS-CoV-2) vaccine booster in patients with chronic liver disease(CLD). A total of 114 patients with CLD who received a SARS-CoV-2 vaccine booster were enrolled in this study. Serum samples were collected from enrolled patients at least 14 days after the booster dose and tested for SARS-CoV-2 neutralizing antibody (novel coronavirus neutralizing antibody, nCoV NTAb) and IgG antibody against SARS-CoV-2 spike binding domain (novel coronavirus spike receptor-binding domain antibody, nCoV S-RBD antibody) levels. The positive rates of nCoV NTAb and nCoV S-RBD in patients with CLD were 87.72% and 91.23%, respectively, after the booster injection of coronavirus disease 2019 (COVID-19) vaccine. The booster injection resulted in the production of nCov NTAb in 66.7% of patients and nCov-SRBD antibody in 71.43% of patients with CLD who failed basic immunization. After basic SARS-CoV-2 immunization, the booster SARS-CoV2 vaccine increased the serum conversion rate and the level of nCov NTAb and nCov-SRBD antibodies in patients with CLD (including patients with cirrhosis). The severity of the liver disease is related to the immune response to COVID-19 vaccine.

## Introduction

In recent years, COVID-19 caused by severe acute respiratory syndrome coronavirus 2 (SARS-CoV-2), has spread worldwide. In order to fight the epidemic, research and development, as well as application of the novel coronavirus vaccine, have been carried out globally ^[1,2]^. SARS-CoV-2 vaccine injection specifically induces a strong protective immune response against COVID-19 ^[2]^. Also, neutralizing antibody levels in vitro indicate a protective effect on COVID-19 ^[3]^. With the passage of time, the antibody level in the human body shows a gradual decrease ^[4]^, requiring people to increase the vaccination to enhance the immune level against the coronavirus and reduce the risk of infection. Since a specific group of people had low immunity, the positive rate of neutralizing antibodies in patients with CLD after two doses of inactivated SARS-CoV-2 vaccine was only 76.8%, which was lower than that of 90.3% in the healthy control group ^[5]^; the positive rate of neutralizing antibodies in decompensated cirrhosis was even lower (66.1–71.6%) ^[6]^. Therefore, patients with CLD need to strengthen their immunity to COVID-19. The safety and immune reactivity of these patients after injection of enhanced SARS-CoV-2 vaccine are a major concern. Thus, the present study aimed to investigate the safety and immunoreactivity of SARS-CoV-2 booster shots in patients with CLD.

## 1. Study design

Patients with CLD who visited the outpatient department of Tianjin Third Central Hospital(Tianjin, China) from July 2021 to June 2022 were included in this study. All subjects received a booster shot of SARS-CoV-2 vaccine. According to the SARS-CoV-2 vaccination guidelines formulated by the National Health Commission of the People’s Republic of China ^[7]^, the booster injection should be given at least 6 months after the previous injection.

### 1.1 Inclusion and exclusion criteria

The inclusion criteria were as follows: 1. Age ≥18 years; 2. Patients with CLD diagnosed clinically or by cases; 3. Receiving SARS-CoV-2 booster shot>14 days; after >; 4. Willing to comply with the study procedures and provide written informed consent.

The exclusion criteria were as follows: pregnancy, lactation, positive COVID-19 or known history of COVID-19 infection, known history of liver transplantation, immunosuppression or immunodeficiency status (including HIV infection), and history of systemic immunosuppression, systemic immunoglobulin, or immune boosters received within 3 months prior to the screening date.

### 1.2 Diagnosis of CLD

#### 1.2.1 Definition of CLD: Liver disease lasting >6 months, including chronic inflammation

(hepatitis B, hepatitis C, non-alcoholic fatty liver disease (NAFLD), alcoholic liver disease (ALD), autoimmune hepatitis (AIH), primary biliary cholangitis(PBC), and primary sclerosing cholangitis(PSC)) with or without cirrhosis.

#### 1.2.2 Diagnostic basis of compensatory cirrhosis (one of the following four) ^[8]^

1. Histology was consistent with the diagnosis of liver cirrhosis;
2. Endoscopy showed esophageal and gastric or ectopic varices of the digestive tract, excluding non-cirrhotic portal hypertension.
3. Imaging examinations such as B-scan ultrasonography, Liver Stiffness Measurement(LSM,)or Computed Tomography(CT)suggest cirrhosis or portal hypertension features: for example, splenomegaly, portal vein ≥1.3 cm, and LSM measurement in line with the diagnostic threshold of cirrhosis of different etiologies.
4. In the absence of histology, endoscopy, or imaging examination, the following abnormalities suggested cirrhosis (2/4 items should be met): ① Platelet (PLT)<100×10^9^/L with no other explanation; ②Serum albumin(ALB)<35 g/L, excluding other causes such as malnutrition or renal disease; ③International Normalized Ratio(INR)>1.3 or prolonged Prothrombin time(PT)(>7days after discontinuation of thrombolytic or anticoagulant drugs); ④Aspartate Aminotransferase (AST)/PLT ratio index (APRI): adult APRI score >2.

#### 1.2.3 Diagnostic basis of decompensated cirrhosis

Based on liver cirrhosis, portal hypertension complications and/or hepatic dysfunction, such as ascites, esophagogastric variceal bleeding, sepsis, hepatic encephalopathy, and hepatorenal syndrome, occur.

### 1.3 Clinical data collection

Basic characteristics (age, gender, nationality, and body mass index (BMI)), liver disease etiology (hepatitis B, hepatitis C, alcoholic liver, and autoimmune liver disease), liver disease severity (CLD and compensated or decompensated cirrhosis), history of medication for liver disease, complications (diabetes, hypertension, arrhythmia, coronary heart disease, and bronchial asthma), blood routine, coagulation function, liver function, COVID-19 antibody test, abdominal ultrasound, liver hardness, and other data were collected.

### 1.4 Safety evaluation of vaccine injection

The data of local and systemic adverse events in patients with CLD were collected within 14 days after SARS-CoV-2 booster injection.

The following adverse reactions were recorded: Local reactions at the injection site and their severity: pain, swelling, redness, induration, and pruritus. Systemic adverse reactions and their severity: fever, dizziness, fatigue, nausea, vomiting, decreased appetite, muscle pain, arthralgia, oropharyngeal pain, cough, allergy, dyspnea, syncope, and pruritus (not at the injection site). Patients may also report other adverse effects not mentioned above. The liver function abnormalities were monitored after booster vaccination.

Abnormal liver function was defined as alanine aminotransferase (ALT, 0–40 U/L),AST (0–40 U/L), γ-glutamyl transpeptidase (GGT, 0–50 U/L), alkaline phosphatase (ALP, 0–150 U/L),ALB(>35 g/L), total bilirubin (TBIL, 0–17.1 μmol /L), and direct bilirubin (DBIL, 0–6 μmol /L) were above the upper limit of normal (ULN).

### 1.5 Assessment of immune response

Serum samples were collected from enrolled subjects at least 14 days after completion of booster injections to quantify the neutralizing antibodies nCoV NTAb and nCoV S-RBD against SARS-CoV-2 by competitive combined chemiluminescence immunoassay (CLIA) (Mindray Bio-Medical Electronics Co., Ltd, Shenzhen, China), according to the manufacturer’s instructions. The assay range was 2.0–400.0 AU/mL, with >10 AU/mL as evidence of immune response and <2.0 U/mL as evidence of no immune response. Subsequently, we defined >10.0 AU/mL as positive and <10.0 AU/mL as negative.

### 1.6 Statistical analysis

Continuous variables were summarized as the median and interquartile range (IQR). Categorical variables were calculated as the percentage of patients in each category. The percentages between the two groups were compared using the chi-square test. Mann-Whitney U (non-parametric) test was used to compare the continuous variables between the two groups. Paired-sample t-tests were used to compare continuous variables between different time points within a group. The univariate and multivariate analyses of factors associated with serological response were performed by fitting binary logistic regression models. Two-sided test significance level was set at p<0.05. Data were analyzed using SPSS software, statistical packages R (http://www.R-project.org, The R Foundation; version 5.0), and EmpowerStats (http://www.empowerstats.com; X&Y Solutions Inc.Boston.MA). The graphs were generated using OriginPro 2022.

### 1.7 Ethics

The study protocol and informed consent were approved by the Ethics Committee of Tianjin Third Central Hospital (Ethics Approval No. IRB2021-027-01). All subjects signed a written informed consent prior to enrollment.

## 2. Results

### 2.1 Characteristics of patients at baseline

A total of 114 patients with CLD who met the inclusion criteria from July 2021 to June 2022 were enrolled in this study. The cohort comprised 62 patients with and 52 patients without cirrhosis; the median age was 52 and 54 years, respectively. Among them, 52 (45.61%) received CoronaVac novel coronavirus vaccine (Sinovac Research & Development Co., Ltd), 46 (40.35%) received BBIBP-CorV vaccine (Beijing Institute of Biological Products), 11 (9.65%) received CanSinoBio Ad5-nCoV vaccine (CanSino Biologics Inc.), and 5 (4.39%) received Zifivax vaccine (CHO cells) (Anhui Zhifei Longcom Biopharmaceutical Co., Ltd). In the cirrhotic and non-cirrhotic groups, hepatitis B virus (HBV) infection accounted for 84.62%(44/52) and 87.10%(54/62) of the causes, of which 60.53%(69/114) received anti-HBV treatment. The most common comorbidities were hypertension, diabetes mellitus, and coronary heart disease (18.42%(21/114), 10.53%(12/114), and 4.39%(5/114), respectively), followed by arrhythmia and asthma. The liver function parameters were normal or stable. Child–Pugh score was at B/C level in 5.88%(3/51) of patients in the cirrhosis group (Table 1).

**Table 1.**
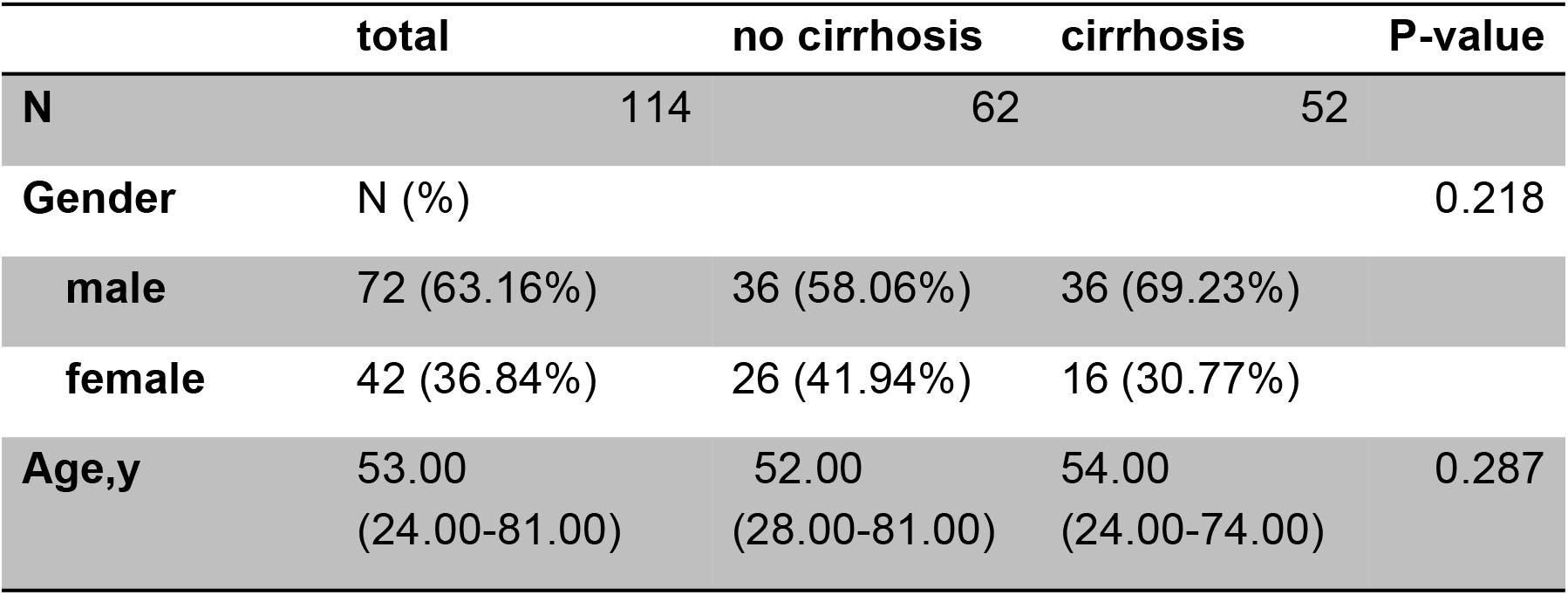
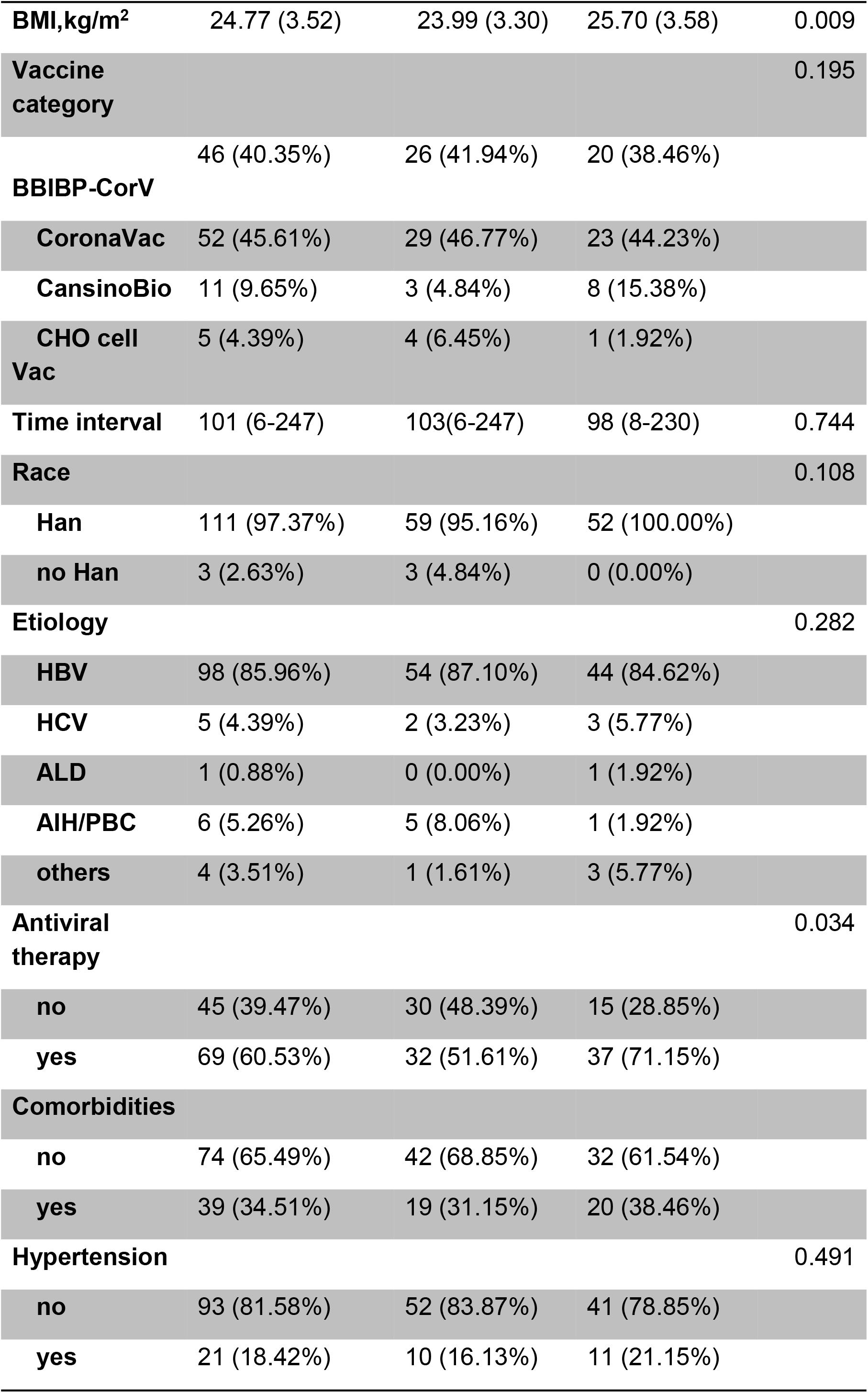
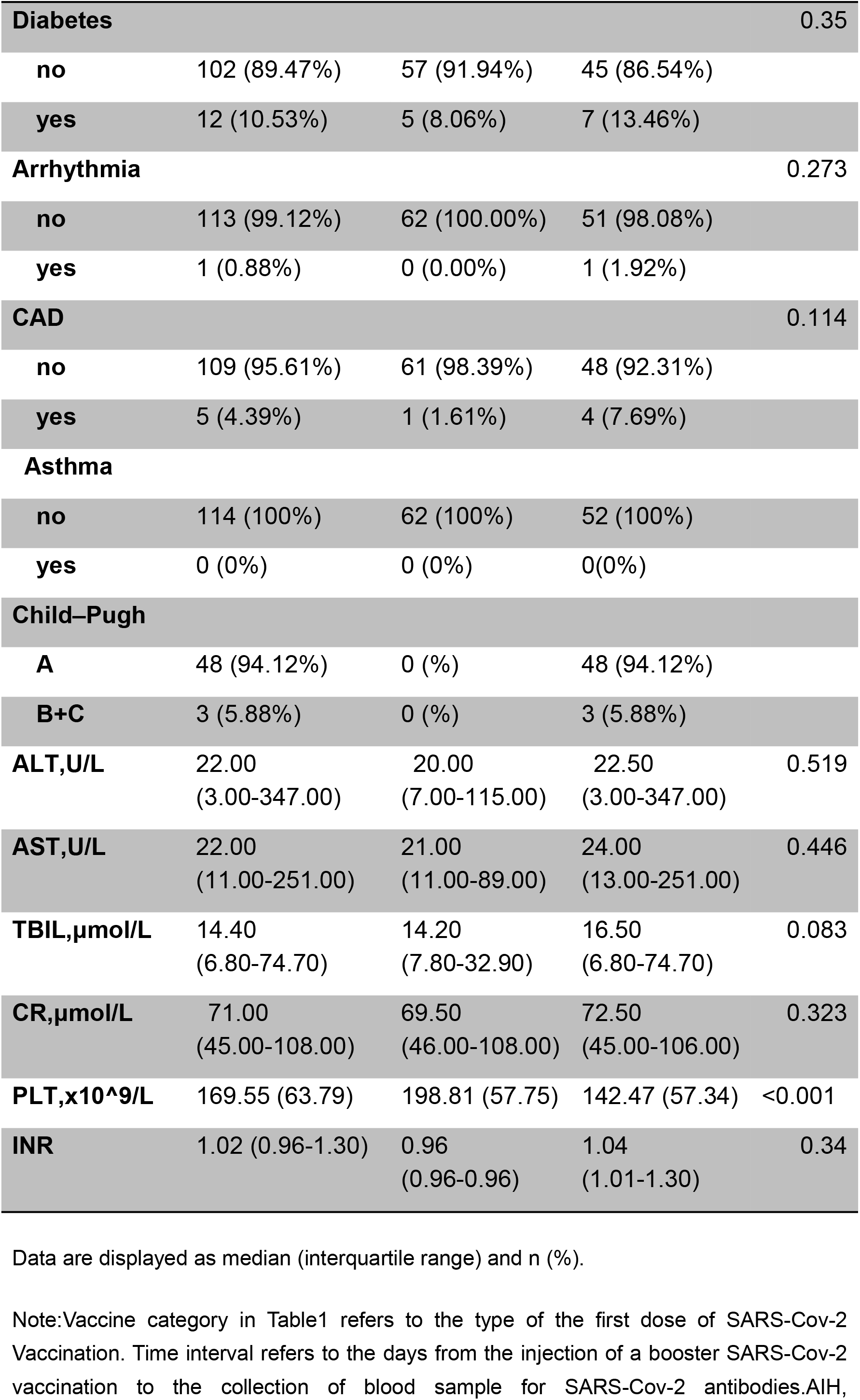
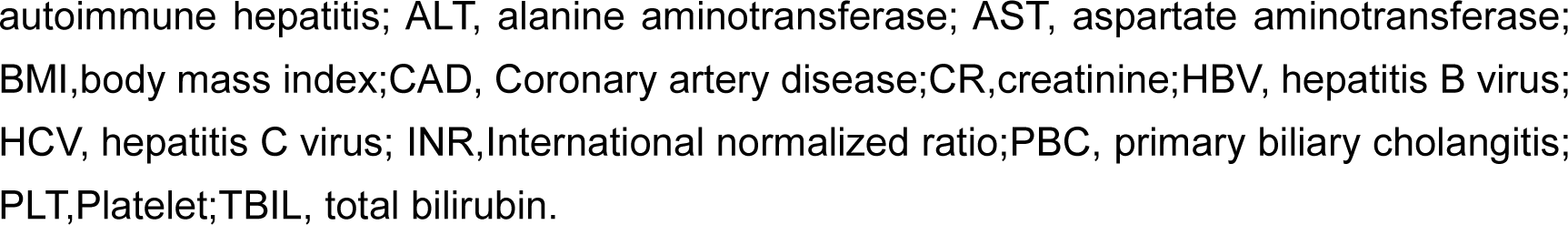
Baseline characteristics of patients with liver disease

### 2.2 Safety evaluation of vaccination

COVID-19 vaccine has a good safety profile in patients with CLD, both cirrhotic and non-cirrhotic groups. A total of 49 adverse reactions occurred in 35/114(30.70%) patients with CLD who received COVID-19 vaccine booster injections.s. Among them, 13/52(25%) patients with cirrhosis and 22/62 (35.48%) patients without cirrhosis reported at least one adverse reaction within 14 days of COVID-19 vaccination, and no significant difference was observed in the adverse reactions between the two groups (p=0.827). The most common local adverse reaction was pain at the injection site, 16/114(14.04%), followed by pruritus, redness, and swelling heat. The most common systemic adverse reactions were fatigue, dizziness, and nausea. Other adverse reactions included drowsiness in 8 cases (1 case with catarrhal symptoms), headache in 2, hiccup with abdominal distension (1 case), diarrhea (1 case), and epigastric pain (1 case); these were mild adverse reactions within grade 2 and were relieved eventually (Table 2).

**Table 2:** Adverse reactions of patients with chronic liver disease after any dose of SARS-CoV-2 vaccines

| characteristics | Total | Cirrhosis | No cirrhosis | P value |
| --- | --- | --- | --- | --- |
|  | (n=114) | (n=52) | (n=62) |  |
| **Local adverse reactions of injection** |  |  |  |  |
| **pain** | 16 (14.04) |  |  | 0.583 |
| **Grade1** | 14 (12.28) | 5 (9.62) | 9 (14.52) |  |
| **Grade2** | 2 (1.75) | 0 (0) | 2 (3.23) |  |
| **itch** | 1 (0.88) |  |  | 0.456 |
| **Grade1** | 1 (0.88) | 1 (1.92) | 0 (0) |  |
| **Swelling** | 0 (0.00) |  |  |  |
| **Grade1** | 0 (0.00) | 0 (0) | 0 (0) |  |
| **turn red** | 0 (0.00) |  |  |  |
| **Grade1** | 0 (0.00) | 0 (0) | 0 (0) |  |
| **Local discomfort** | 1 (0.88) |  |  | 0.456 |
| **Grade1** | 1 (0.88) | 1 (1.92) | 0 (0) |  |
| **Systemic adverse reactions** |  |  |  |  |
| **fever** | 1 (0.88) |  |  | 0.544 |
| **Grade1** | 1 (0.88) | 0 (0) | 1 (1.61) |  |
| **dizzy** | 3 (2.63) |  |  | 0.249 |
| **Grade1** | 3 (2.63) | 0 (0) | 3 (4.84) |  |
| **weakness** | 5 (4.39) |  |  | 0.583 |
| **Grade1** | 5 (4.39) | 2 (3.85) | 3 (4.84) |  |
| **Myalgia** | 1 ( 0.88) |  |  | 0.544 |
| **Grade1** | 1 (0.88) | 0 (0) | 1 (1.61) |  |
| **Arthralgia** | 1 (0.88) |  |  | 0.544 |
| **Grade1** | 1 (0.88) | 0 (0) | 1 (1.61) |  |
| **cough** | 1 (0.88) |  |  | 0.544 |
| **Grade1** | 1 (0.88) | 0 (0) | 1 (1.61) |  |
| **nausea** | 4 (3.51) |  |  | 0.622 |
| **Grade1** | 4 (3.51) | 2 (3.85) | 2 (3.23) |  |
| **Anorexia** | 2 (1.75) |  |  | 0.499 |
| **Grade1** | 2 (1.75) | 0 (0) | 2 (3.23) |  |
| **others** | 13 (11.40) |  |  | 0.376 |
| **Grade1** | 13 (11.40) | 4 (7.69) | 9 (14.52) |  |
| **Total adverse reactions cases** | 35(30.70) | 13 (25.00) | 22 (35.48) | 0.827 |
| **Grade1** | 34 (97.14) | 13 (100) | 21 (95.45) |  |
| **Grade2** | 1 (2.86) | 0 (0) | 1 (4.55) |  |
| **Total adverse reactions times** | 49 | 15 | 34 | 0.342 |
| **Grade1** | 47 (95.92) | 15 (100) | 32 (94.12) |  |
| **Grade2** | 2 (4.08) |  | 2 (5.88) |  |

Data are displayed as n (%).
Note: Grade 1 of pain at injection site: slight pain. Grade 2: Moderate pain, affecting daily household activities. Grade 3: severe pain, affecting self-care activities of daily living.

Of the 84 patients with available laboratory data for follow-up, 2 (2.38%) reported grade 2 ALT elevation (defined as 2 ULN<ALT≤5 ULN), 2 (2.38%) reported grade 3 ALT elevation (defined as ALT>5 ULN), and 1 (1.19%) reported grade 2 AST elevation (defined as 2 ULN<AST≤5 ULN). These participants showed an improvement with relevant treatment.

### 2.3 Immune response evaluation

#### 2.3.1 Changes in nCoV NTAb and nCoV S-RBD antibody levels after booster vaccination in patients with CLD

In this study, among 114 patients with liver disease after receiving the booster shot of COVID-19 vaccine, 100(87.72%) patients were positive for nCoV NTAb%. A total of 104(91.23%) patients were positive for nCoV S-RBD antibody. The positive rates of nCoV NTAb and nCoV S-RBD antibodies in the non-cirrhotic and cirrhotic groups were 93.55%(58/62), 80.77%(42/52) and 98.39%(61/62), 82.69%(43/52), respectively. The positive rates of both neutralizing antibodies in the non-cirrhotic group were significantly higher than those in the cirrhotic group (p<0.05). The median levels of neutralizing antibodies were 31.98 (5.72-1024.00)AU/mL for nCoV NTAb and 112.65 (1.91-1001.00)AU/mL for nCoV S-RBD, and the median levels of nCoV NTAb antibodies in the non-cirrhotic and cirrhotic groups were 31.41(7.55-401.00) AU/mL and 32.64 (5.72-1024.00)AU/mL, respectively, and those of nCoV S-RBD were 102.94(3.00-1001.00)AU/mL and 119.41(1.91-1000.00)AU/mL, respectively (Table 3, Figure 1).

**Table 3.**
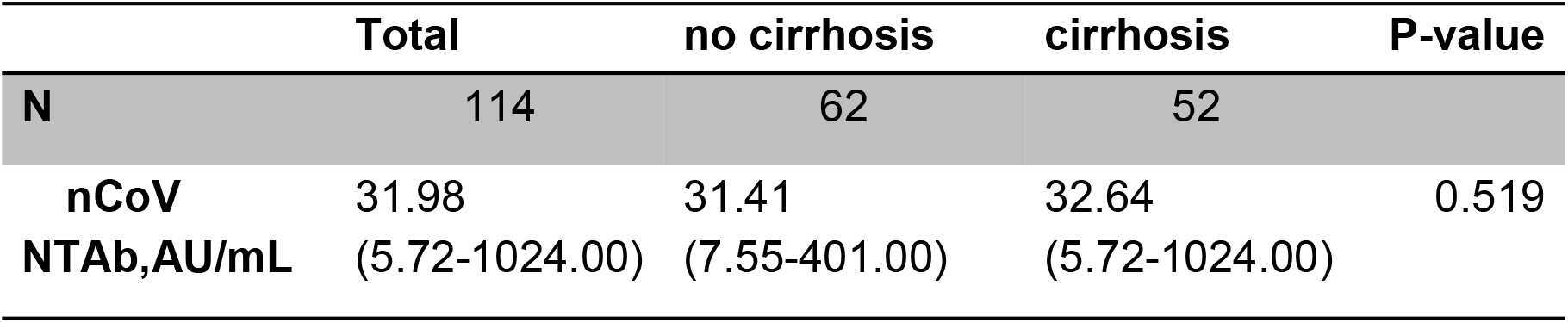
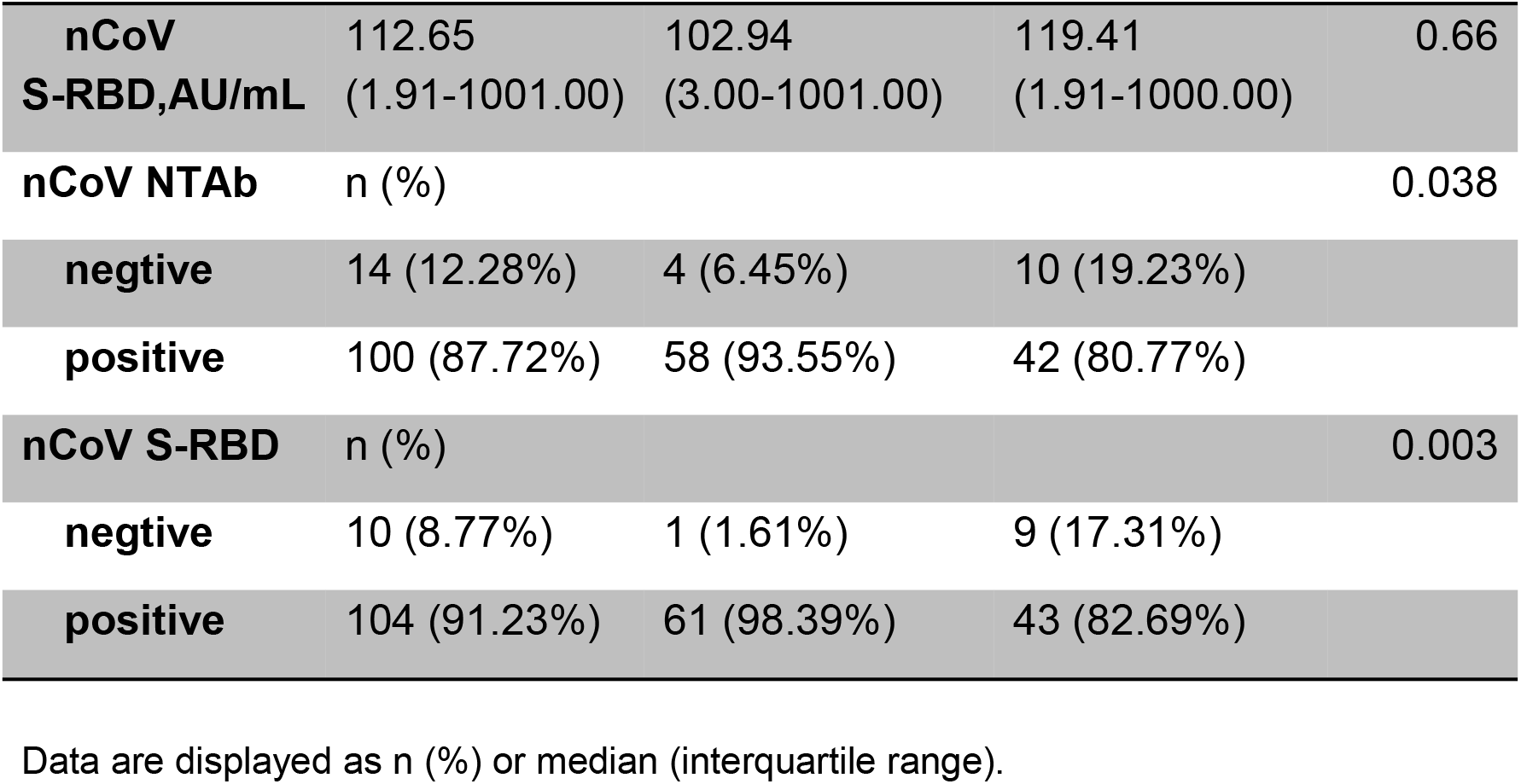
Evaluation of the immunogenicity of a booster SARS-Cov-2 vaccination

**Figure 1.**
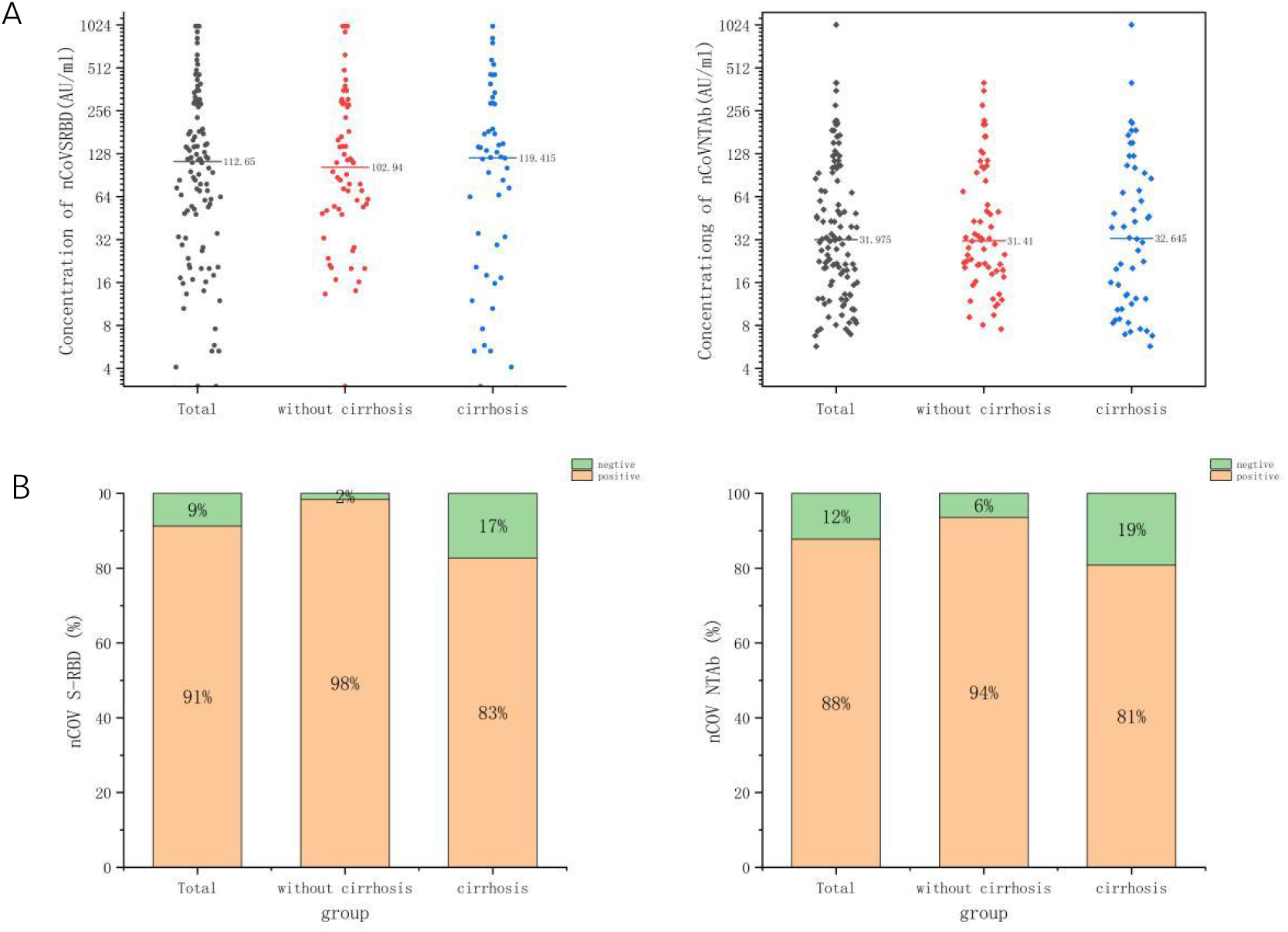
Serological response of a booster SARS-CoV-2 vaccines in patients with chronic liver diseases. Concentrations(A) and positive rates (B)and of nCoV S-RBD and nCoV NTAb induced after a booster SARS-CoV-2 vaccines in all participants, participants without cirrhosis, participants with cirrhosis. Concentrations over 10.0 AU/mL were considered as positive, and concentrations below 10.0 AU/mL as negative.

#### 2.3.2 Dynamic changes in immune response in patients with CLD after basic immunization and booster injection

Among the patients with CLD included in this study, vaccine antibody levels were detected in 22 patients after basic immunization and booster injection of COVID-19 vaccine. The dynamic analysis of the antibody levels in these 22 patients showed that the positive rate of NTAB antibody increased from 59.09%(13/22) to 86.36%,(19/22) and the positive rate of SRBD antibody increased from 68.18%(15/22) to 90.91%(20/22) after booster injection of COVID-19 vaccine. The antibody level of patients with the CLD increased significantly after the COVID-19 vaccine booster. The median antibody level of nCov-NTAb increased significantly from 11.24 (4.41–38.26) to 59.14 (5.72–279.38) AU/mL in 22 patients with CLD, with a mean increase of 5.26-fold, while the median antibody level of nCov-SRBD increased from 27.27 (2.90–169.47) to 219.10 (2.55–579.46) AU/mL, with a mean increase of 8.03-fold. The difference was statistically significant (p<0.01). Among the patients with CLD who failed basal immunization, the booster injection developed neutralizing antibodies in 66.7% (6/9) and anti-S-RBD antibodies in 71.43% (5/7), which is crucial to improving their immune response (Table 4, Figures 2 and 3).

**Table 4.** Comparison of vaccine antibody levels before and after booster injection of COVID-19 vaccine in 22 patients

|  | **Basic immunization vaccine injection** | **Booster shot** | **P value** |
| --- | --- | --- | --- |
| **n** | 22 | 22 |  |
| **nCov NTAbs,AU/mL** | 11.24 (4.41-38.26) | 59.14 (5.72-279.38) | <0.001 |
| **nCov S-RBD,AU/mL** | 27.27<br>(2.90-169.47) | 219.10<br>(2.55-579.46) | <0.001 |
| **nCov NTAbs** | n (%) |  | 0.042 |
| **negative** | 9 (40.91%) | 3 (13.64%) |  |
| **positive** | 13 (59.09%) | 19 (86.36%) |  |
| **nCov S-RBD** | n (%) |  | 0.062 |
| **negative** | 7 (31.82%) | 2 (9.09%) |  |
| **positive** | 15 (68.18%) | 20 (90.91%) |  |

Data are displayed as n (%) or median (interquartile range).

**Figure 2.**
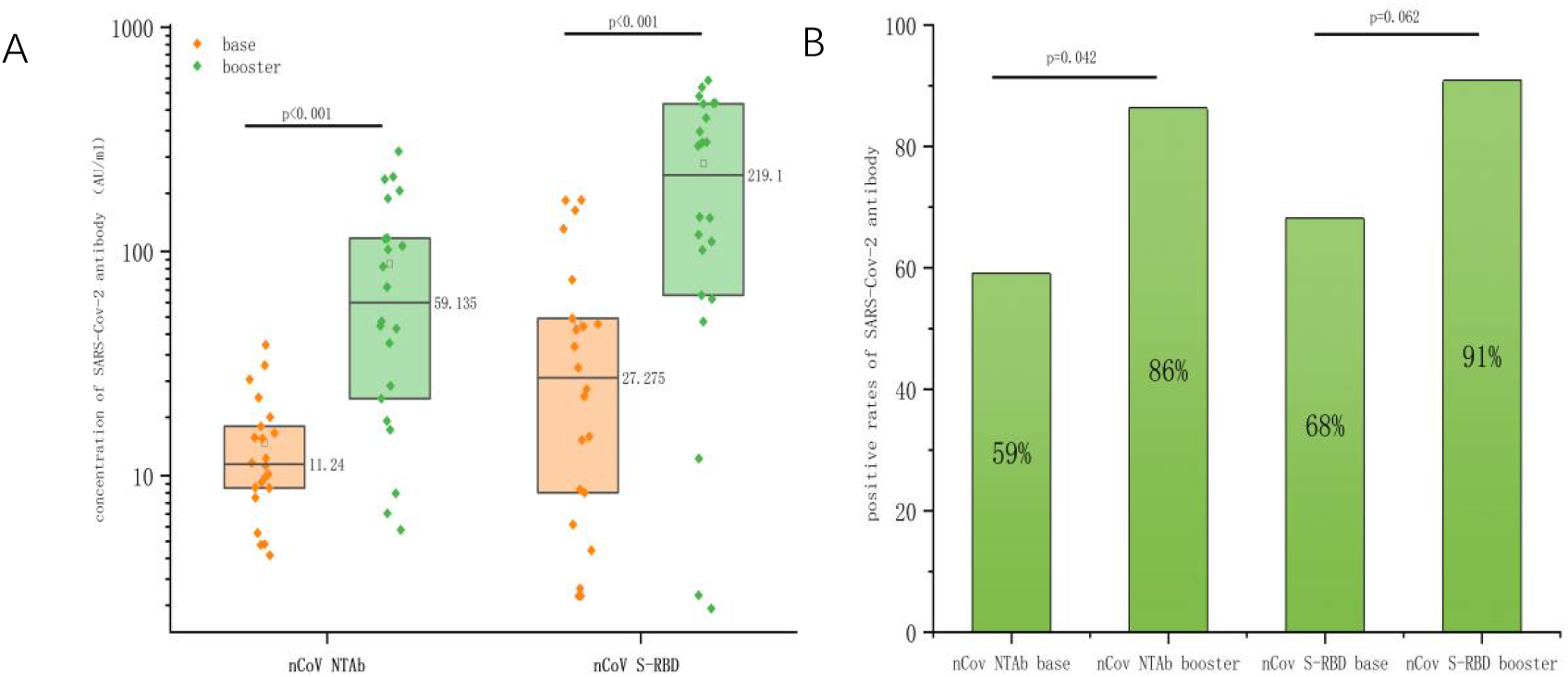
Comparison of SARS-Cov-2 antibody levels before and after booster injection of COVID-19 vaccine. Concentrations(A) and positive rates (B) of nCoV NTAb and nCoV S-RBD after the base and the booster injection of SARS-CoV-2 vaccine in 22 patients.

**Figure 3.**
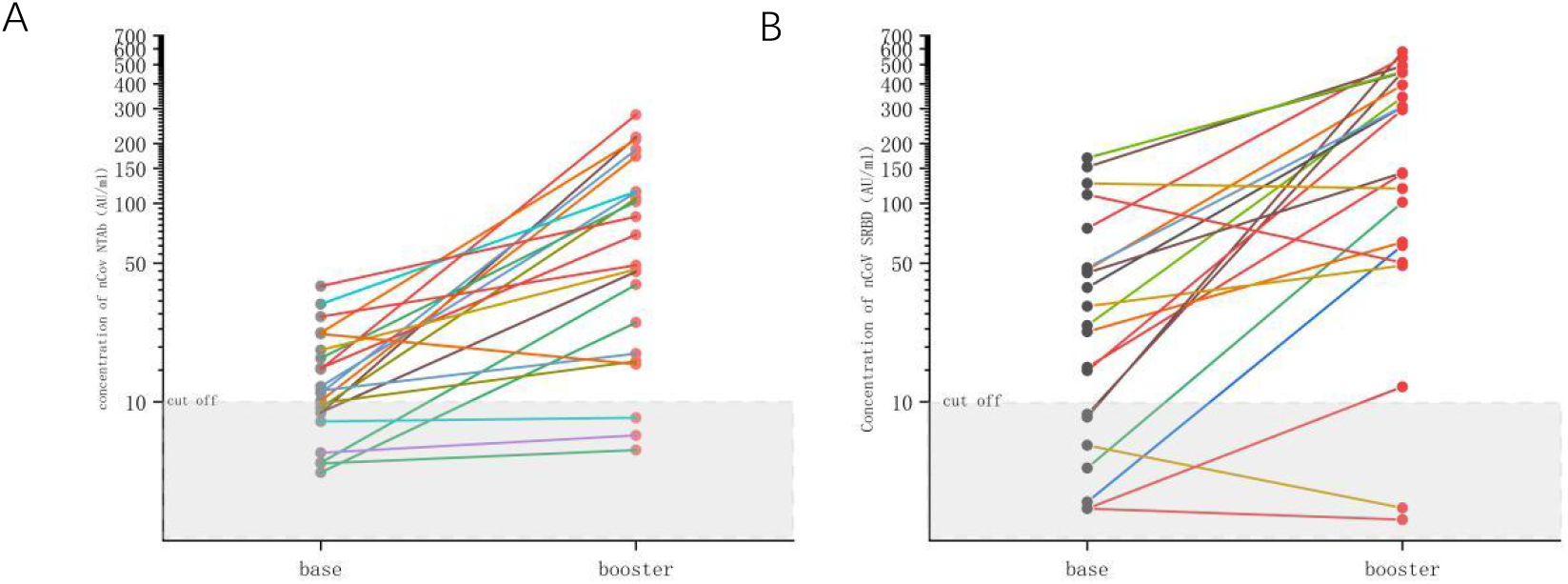
Comparison of SARS-Cov-2 antibody levels before and after booster injection of COVID-19 vaccine. There were 22 patients who tested the SARS-Cov-2 antibody after completing both the basic immunization and booster injection of COVID-19 vaccine.The change of concentration of nCoV NTAb (A) and nCoV S-RBD (B) were show in figure.Concentrations over 10.0 AU/mL were considered as positive, and concentrations below 10.0 AU/mL as negative. Of patients who failed basal immunization, the booster SARS-Cov-2 vaccination can make 6 of 9 patients with negative nCoV NTAb results turn to be positive,5 of 7 patients with negative nCoV S-RBD result turn to be positive.

2.3.3 Logistic binary regression analysis of potential correlations of serological response after COVID-19 vaccine booster in all patients did not show any significant correlation between antibody level and baseline status in patients with CLD. The baseline indicators, such as gender, age, height, weight, BMI, etiology, comorbidities, and baseline liver function, may or may not be associated with baseline cirrhosis. Compared to non-cirrhotic patients, the positive rates of nCov NTAb and nCoV S-RBD antibodies were significantly lower in cirrhotic patients (Table 5).

**Table 5.**
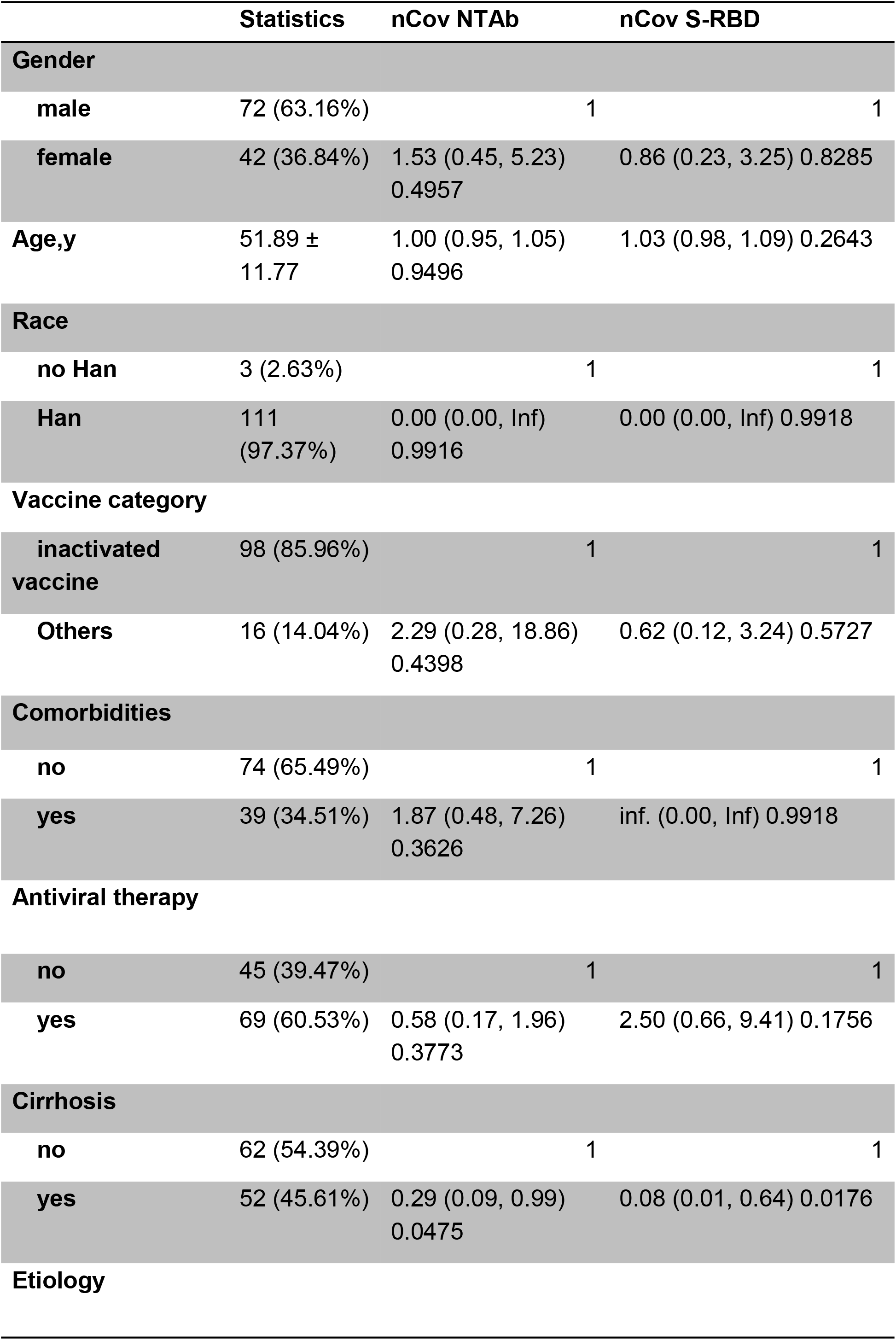
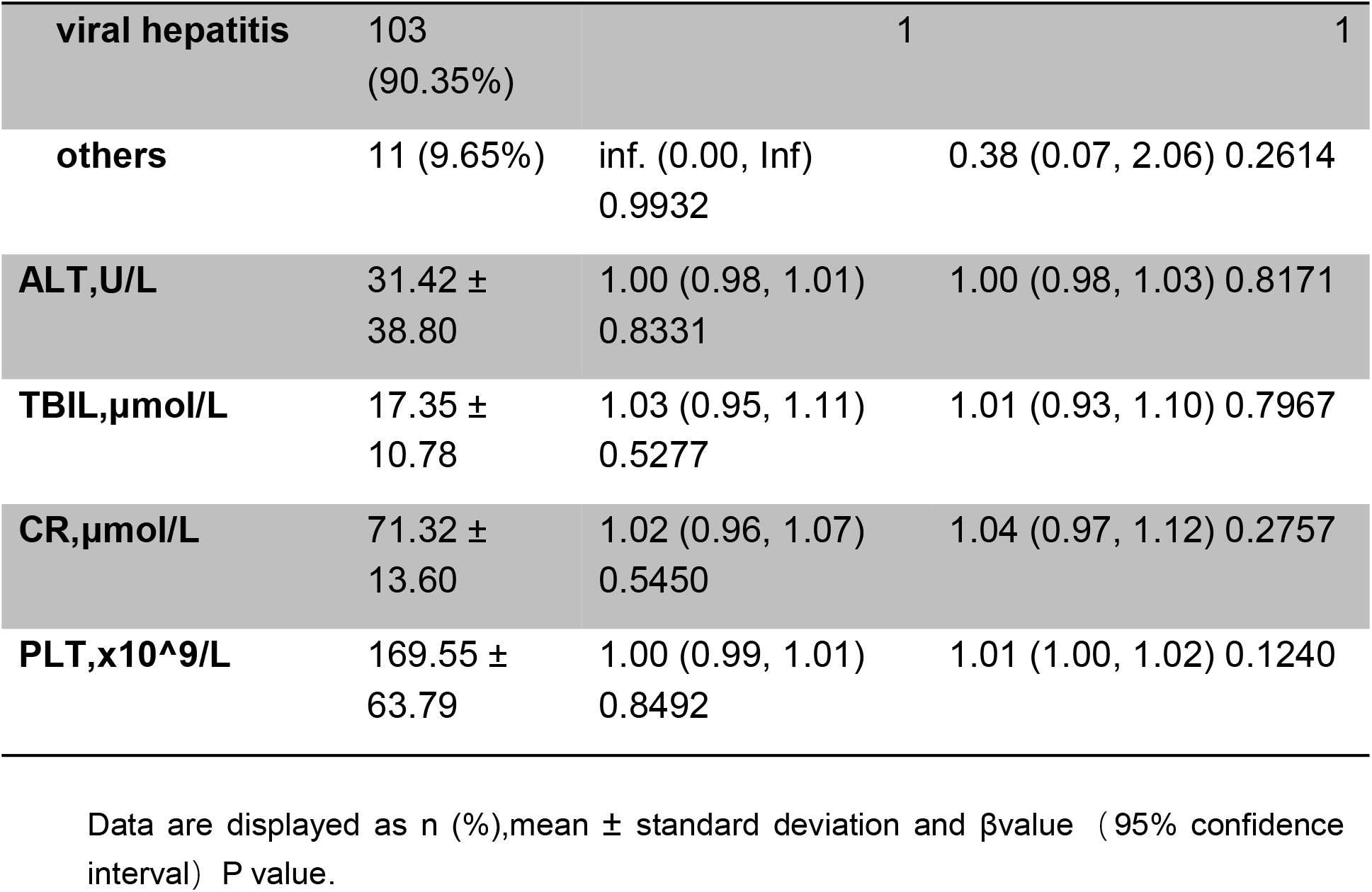
Factors for serological response to SARS-CoV-2 vaccination in patients with chronic liver disease

## 3. Discussion

In December 2019, the global situation of the COVID-19 epidemic was still grim, rendering the elderly and people with underlying diseases prone to infection and serious consequences. Vaccine is a key cost-effective tool for controlling the COVID-19 pandemic, and COVID-19 vaccine injection has reduced the hospitalization rate, severe illness rate, and death rate of COVID-19 patients ^[9,10]^. Neutralizing antibody levels decay over time ^[4,11]^, and most populations require booster shots to increase the antibody levels in order to maintain immunity against COVID-19. Ai et al. showed that after receiving two doses of whole virion-inactivated “starter” vaccine, the third protein subunit vaccine heterologous booster was safe for healthy adults; it is highly immunogenic, which significantly evokes and enhances the immune response to SARS-CoV-2 and its variants ^[12]^. Cao et al. evaluated the immunogenicity in healthy volunteers who received two doses of CoronaVac inactivated vaccine, followed by an independent injection of either the original inactivated CoronaVac vaccine or a booster of the recombinant protein vaccine ZF2001; consequently, the booster injection rapidly induced the high levels of humoral immunogenicity ^[13]^. For individuals who received booster vaccines more than 5 months apart, the third dose resulted in a 5–7-fold rise in serum neutralization titers ^[14,15]^. Therefore, boosting immunization with COVID-19 vaccine is a good strategy to combat the global COVID-19 epidemic.

The immune function of patients with CLD is impaired. Previous studies have confirmed that the immune response of patients with CLD to basal SARS-COV-2 vaccine is lower than that of healthy populations ^[5,16,17]^. Therefore, this group of patients may have a greater need for booster injections. The literature on booster doses of SARS-COV-2 vaccine in patients with CLD is minimal. Davidov et al. assessed immune responses in liver transplant recipients before and after the third dose of the BNT162b2mRNA vaccine and found that enhanced vaccination significantly increased the humoral immune response rate (56% *vs*. 98%) and the level of anti-S-RBD IgG and neutralizing antibodies ^[18]^. Another study by Hartl et al. confirmed a significant increase in the median S-RBD antibody levels in patients with autoimmune hepatitis (AIH) after the third COVID-19 vaccination ^[19]^. However, these studies only included patients with liver transplantation and AIH, excluding patients with CLD, especially cirrhosis.

The results of our single-center study showed that patients with CLD had an overall neutralizing antibody positivity rate of 87.72%, nCoV S-RBD antibody positivity rate of 91.23%, nCoV NTAb level of 31.98 AU/mL, and nCoV S-RBD antibody level of 112.65 AU/mL after receiving the booster shot of COVID-19 vaccine. However, previous studies have shown that the positive rate of nCoV NTAb in patients with CLD after receiving two doses of SARS-CoV-2 vaccine was only 76.8%, and the level of neutralizing antibodies was 17.7AU/mL ^[5,20]^. These levels suggested that the third dose of COVID-19 vaccine significantly improved the antibody seroconversion rates and increased the absolute levels of neutralizing antibodies in patients with CLD. In the present study, 22 patients were continuously tested for nCoV NTAb and nCoV S-RBD antibodies levels after completing the basic immunization and booster injection of COVID-19 vaccine. The results were analyzed dynamically; the nCoV NTAb positivity rate increased from 59.09% to 86.36% and the nCoV S-RBD IgG antibody positivity rate increased from 68.18% to 90.91% in patients with CLD after the booster injection. The median nCov NTAb level increased significantly from 11.24 AU/mL to 59.14 AU/mL in 22 patients after the booster vaccination; the median S-RBD IgG antibody level increased significantly from 27.27 AU/mL to 219.10 AU/mL. The booster injection resulted in the production of nCoV NTAb and nCoV SRBD antibodies in 66.7% and 71.43% of patients with CLD who failed basal immunization, respectively. The findings suggested that booster shots are essential to improving the immune response in patients with CLD, especially in those with failed basal immunity.

Previous studies have reported cases of patients with cirrhosis who were infected with COVID-19 even after receiving the vaccine ^[21]^. After 28 days of receiving mRNA-based COVID-19 vaccine, the virus infection rate of cirrhotic patients was 66.8% lower than that of unvaccinated cirrhotic patients and much lower than that of the general population ^[22]^. In the present study, 52/114 patients with CLD were cirrhotic. The booster vaccination also improved the antibody seroconversion rate in cirrhotic patients, in which the nCoV NTAb and nCoV S-RBD antibodies positivity rates were 80.77% and 82.69%, respectively, but lower than those in non-cirrhotic patients with CLD. Albillos et al. pointed out that the weakened response to vaccination in patients with cirrhosis may be related to the immune dysfunction of patients with cirrhosis ^[23]^. As an immune and endocrine organ, the liver is responsible for a variety of metabolic and immune processes in the body. The final clearance of the virus in COVID-19 patients requires a cellular immune response involving T cells ^[24]^ and the participation of the liver, while in patients with CLD, the cellular immune response is weakened due to impaired liver function. Another study has shown that patients with severe liver disease have a diminished humoral immune response to SARS-CoV-2 vaccine ^[25]^. In the present study, the antibody positivity rate was lower in the cirrhotic group after receiving the booster shot of COVID-19 vaccine compared to the non-cirrhotic group. Multivariate analysis showed that cirrhosis is an independent risk factor affecting the antibody seroconversion rate after vaccination, which supports the theory that the severity of the liver disease affects the immune response to the vaccine, thereby indicating that additional doses of vaccine booster are required to provide effective immune protection in patients with cirrhosis.

Furthermore, the rate of adverse reactions in patients with CLD was low after the injection of COVID-19 vaccine booster shot, and the adverse reactions were mild and common, such as local pain, dizziness, and fatigue, which were relieved eventually. Previous studies have also confirmed that generally, patients with cirrhosis are tolerant to COVID-19 vaccine, and the most common local adverse reaction is pain at the injection site; all these local adverse reactions can resolve spontaneously ^[20,25]^. Thus, the COVID-19 vaccine can be safely injected into patients during the stable period of their CLD. Considering the acceptable risk of the adverse effects of vaccination and the potential for better immune protection, patients with CLD, especially those with cirrhosis, should be actively vaccinated with booster vaccines.

To the best of our knowledge, this is the first study focusing on patients with CLD receiving a booster dose of the COVID-19 vaccine. Since this is a real-world study, patients with CLD who did not receive a booster shot of COVID-19 vaccine and healthy individuals were not included as controls. Among the subjects included in this study, the number of cases with continuous dynamic antibody levels was small, and the results need to be substantiated with additional data. Also, the liver in the majority of patients was chronic hepatitis B, while only a few had other etiologies; therefore, studies focused on other types and different states of liver diseases are needed in the future. In addition, this was a single-center study; most of the subjects were from the same geographical area, and most received inactivated vaccines. Thus, in future studies, the sample size should be expanded to include more regional populations and diverse varieties of COVID-19 vaccine booster shots to validate our findings.

In conclusion, the current study showed that after receiving basal COVID-19 vaccination, the booster SARS-CoV2 vaccine significantly improved the nCoV NTAb and nCoV S-RBD antibodies seroconversion rates and antibody levels in patients with CLD, including those with mild and acceptable adverse effects. These findings strongly support the importance of booster vaccination for patients with CLD after completion of the standard vaccination regimen, especially in those with failed underlying immunization.

## Data Availability

All data produced in the present study are available upon reasonable request to the authors.

## 5. Data availability statement

The data that support the findings of this study are available from the corresponding author on reasonable request.

